# The Expertise Paradox: Who Benefits from LLM-Assisted Brain MRI Differential Diagnosis?

**DOI:** 10.1101/2025.10.28.25338816

**Authors:** Severin Schramm, Bastien Le Guellec, Marlene Topka, Mortimer Svec, Paul Backhaus, Viktor Maria Eisenkolb, Evamaria O. Riedel, Mirjam Beyrle, Paul-Sören Platzek, Constanze Ramschütz, Karolin J. Paprottka, Martin Renz, Jannis Bodden, Jan S. Kirschke, Sebastian Ziegelmayer, Felix Busch, Marcus R. Makowski, Lisa Adams, Keno Bressem, Dennis M. Hedderich, Benedikt Wiestler, Su Hwan Kim

**Author notes:** Corresponding Author: Su Hwan Kim, MD MSc.

## Abstract

**Purpose:** To evaluate how reader experience influences the diagnostic benefit from LLM assistance in brain MRI differential diagnosis.

**Materials and Methods:** Neuroradiologists (n = 4), radiology residents (n = 4), and neurology/neurosurgery residents (n = 4) were recruited. A dataset of complex brain MRI cases was curated from the local imaging database (n = 40). For each case, readers provided a textual description of the main imaging finding and their top three differential diagnoses (“Unassisted”). Three state-of-the-art large language models (GPT-4.1, Gemini 2.5 Pro, DeepSeek-R1) were prompted to generate top-three differentials based on the clinical case description and reader-specific findings. Readers then revised their differential diagnoses after reviewing GPT-4.1 suggestions (“Assisted”). To evaluate the association between reader experience and diagnostic benefit, a cumulative link mixed model (CLMM) was fitted, with change in diagnostic result as ordinal outcome, reader experience as predictor, and random intercepts for rater and case.

**Results:** LLM-generated differential diagnoses achieved the highest top-3 accuracy when provided with image descriptions from neuroradiologists (top-3: 78.8-83.8%), followed by radiology residents (top-3: 71.8-77.6%), and neurology/neurosurgery residents (top-3: 62.6-64.5%). In contrast, mean relative gains in top-3 accuracy through LLM assistance diminished with increasing experience, with +19.2% for neurology/neurosurgery residents (from 43.2% to 62.6%), +14.7% for radiology residents (from 59.6% to 74.4%), and +4.4% for neuroradiologists (from 83.1% to 87.5%). The CLMM demonstrated a significant negative association between reader experience and diagnostic benefit from LLM assistance (β = −0.10, p = 0.005).

**Conclusion:** With increasing reader experience, absolute diagnostic LLM performance with reader-generated input improved, while relative diagnostic gains through LLM assistance paradoxically diminished. Our findings call attention to the divergence between standalone LLM performance and clinically relevant reader benefit, and emphasize the need to account for human-AI interaction in this context.

## Introduction

In recent years, various applications of large language models (LLMs) in radiology have been demonstrated, including study protocoling [1, 2], the generation of an impression section of a radiology report [3, 4], and data extraction from free-text reports [5–7]. In addition, numerous studies have investigated the potential of LLMs in radiological differential diagnosis, either as standalone tools generating diagnoses from textual imaging findings and key images [8–11], or as assistive tools offering diagnostic suggestions for consideration by human readers [12–15]. Potentially owing to heterogeneity in reader cohorts and study designs, this latter group of studies produced mixed results, with some studies reporting improved accuracy [12, 13] while others did not [14, 15].

Importantly, human-AI interaction is a crucial factor influencing the diagnostic benefit readers can derive from LLM assistance. A recent large-scale investigation found that while LLM assistance enhanced the diagnostic accuracy of board-certified internal medicine physicians based on clinical case vignettes, the same LLM alone remained superior, indicating that the human experts frequently rejected even correct LLM suggestions [16]. Similarly, radiology residents were reported to reject correct LLM-suggested diagnoses in 17.9% of cases [12], underlining the critical challenge of effectively validating LLM outputs. In addition, the diagnostic performance of LLMs is mediated by the quality of its inputs. Radiologist-generated textual imaging findings were found to be the primary determinant of LLM accuracy [17], indicating that LLM performance may fluctuate substantially with variation in reader inputs.

Against this background, we aimed to investigate how reader experience influences the diagnostic benefit from LLM assistance in brain MRI differential diagnosis. We hypothesized that expert-written imaging findings would better encode discriminative features and yield more accurate LLM-generated differential diagnoses than those of novice readers, but that the diagnostic benefit of LLM assistance would be offset by the higher baseline performance of expert readers.

## Methods

This study was approved by ethics committee of the Technical University of Munich, and the need for informed consent was waived.

### Dataset

40 brain MRI cases from the local imaging database were included. These cases were selected from a local neuroradiology case collection (n = 778). Inclusion criteria were a confirmed diagnosis, established either histopathologically or by independent consensus of at least two neuroradiologists, based on all available clinical and follow-up information. Additionally, each case had to be of sufficient diagnostic complexity to be considered appropriate for use in neuroradiology subspecialty examinations, as determined by two board-certified neuroradiologists (B.W. and D.M.H., each with 10 years of experience). Exclusion criteria included imaging of a modality or anatomical region other than brain MRI, inadequate image quality, or an unconfirmed diagnosis. Conditions spanned the entire neuroradiological spectrum, including vascular (e.g. Moyamoya syndrome), infectious (e.g. toxoplasmosis), neoplastic (e.g. chondrosarcoma), degenerative (e.g. Huntington’s disease), metabolic (e.g. Wernicke encephalopathy), demyelinating (e.g. progressive multifocal leukoencephalopathy), traumatic (e.g. diffuse axonal injury), and developmental (focal cortical dysplasia) disorders. Scans were obtained between January 1, 2009, and April 30, 2024. All cases have been published previously [17]. Yet, the clinical case descriptions were published only in January 2025, after the knowledge cutoff date of all models used in this study, thereby excluding the possibility of performance overestimation due to data leakage. To prevent potential bias, readers were instructed a priori to report any cases they recognized from prior clinical involvement, and these were excluded from the analysis to ensure that all remaining cases were unfamiliar to the readers. An overview of clinical cases with respective medical histories is provided in Supplement 1.

### Study Design

An overview of the study design is shown in Figure 1. Neuroradiologists (n = 4), radiology residents (n = 4), as well as neurology and neurosurgery residents (hereinafter referred to as “NL/NS residents”, n = 4; n = 2 each) were recruited from a single academic center. Based on the MRI scan, a condensed medical history and patient demographics, readers created a textual description for the main image finding, which was clearly annotated with one or more arrows. In addition, readers indicated a ranked list of up to three differential diagnoses, along with an overall confidence score on a 5-point Likert scale. Brain MRI images were accessed using the local PACS system. Textual MRI findings were then provided to three LLMs (Gemini 2.5 Pro, GPT-4.1, DeepSeek-R1) which were prompted to equally generate the top-3 differential diagnoses. Subsequently, readers reviewed the differential diagnoses of one representative LLM (GPT-4.1), generated from their own textual findings, and provided their final top-three diagnoses and confidence level taking into consideration the model’s suggestions and explanations. GPT-4.1 was selected at random after a preliminary analysis demonstrated comparable diagnostic accuracy among the three tested models. Cases were presented to the readers in a randomized order, which was identical across the unassisted and LLM-assisted reading conditions for each reader. To avoid additional confounding factors from external research, readers were instructed to refrain from consulting textbooks or online resources and to assess the cases solely based on their existing knowledge. Primary outcome was case-level change in top-3 accuracy from unassisted to assisted reading. Secondary outcomes included correctness and completeness ratings of reader-generated imaging findings.

**Figure 1:**
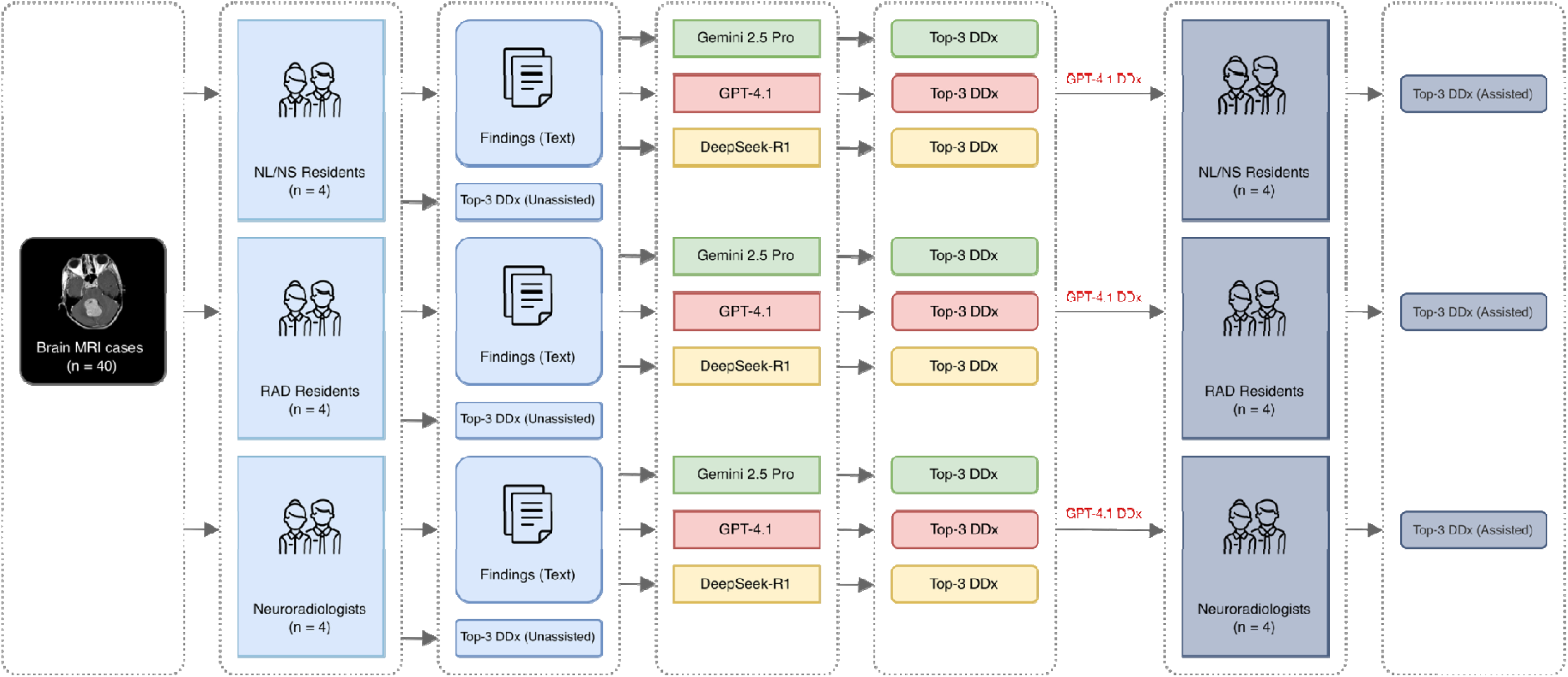
Study Design. Readers first generated a textual description of the main imaging finding and provided their top three differential diagnoses (unassisted). Gemini 2.5 Pro, GPT-4.1, and DeepSeek-R1 were then prompted to generate their top three differential diagnoses based on a condensed medical history and each reader’s finding description. Subsequently, readers reviewed GPT-4.1’s differential diagnoses derived from their own descriptions and provided their final, integrated top three differential diagnoses (assisted). Icons were obtained from flaticon.com. Sample brain MRI image was obtained from: https://doi.org/10.53347/rID-10047. N: Neurology. NS: Neurosurgery. RAD: Radiology. DDx: differential diagnosis.

### LLM Setup

Gemini 2.5 Pro (“gemini-2.5-pro-preview-03-25”, proprietary model; Google DeepMind, London, UK), GPT-4.1 (“gpt-4.1-2025-04-14”, proprietary model; OpenAI, Inc., San Francisco, USA), and DeepSeek-R1 (“deepseek-r1”, open-source; Hangzhou DeepSeek Artificial Intelligence Basic Technology Research Co., Ltd., Hangzhou, China) were used as representative state-of-the-art LLMs at the time of the study. Gemini 2.5 Pro and GPT-4.1 were accessed through the official APIs of Google (https://ai.google.dev/api) and OpenAI (https://platform.openai.com/docs/models), respectively. DeepSeek-R1 was accessed via Fireworks AI (https://fireworks.ai/models), a generative AI inference platform with servers located in the United States and Europe. Models were prompted as follows:

*“You are an experienced neuroradiologist. Below is a case presentation including patient demographics, relevant clinical history, and brain MRI findings. Based on this information, provide your top three differential diagnoses ranked in order of likelihood*.

*CASE: {case_description}*

*For each differential diagnosis, briefly explain your reasoning.”*

The imaging findings were presented to the LLM in German, the native language of the readers. No local fine-tuning or modification was performed. To ensure deterministic outputs, the temperature hyperparameter was set to 0 for all three models. No random seeds beyond the default API configurations were applied. All three models were configured to generate outputs in structured JSON format to ensure consistent and machine-readable responses.

Queries were performed on 3 May 2025. Model outputs were provided to readers without any post-processing.

### Assessment of Diagnostic Accuracy and Image Descriptions

In general, only responses indicating the exact pathologic entity were counted as correct (e.g. “Alzheimer’s disease” was counted as incorrect in a case of posterior cortical atrophy). Edge cases in which the response indicated a correct but less specific diagnosis or related but not identical diagnosis were adjudicated by two board-certified neuroradiologists in consensus (B.W. and D.M.H.) (Supplement 2).

A board-certified neuroradiologist with six years of experience (B.L.G.), who did not participate as a reader and was blinded to the readers’ experience levels but had access to the reference diagnoses and additional online resources (e.g., www.radiopaedia.org), rated the imaging descriptions for correctness and completeness using a 4-point Likert scale (see Supplement 3 for rating criteria).

Top-3 diagnostic accuracy by reader group (proportion of cases where the correct diagnosis was included in the top 3 diagnoses) was determined for unassisted readers, LLMs with reader input, and GPT-4.1-assisted readers. Additionally, the diagnostic benefit of readers was determined by subtracting top-3 accuracy of unassisted readers from top-3 accuracy of assisted readers.

### Statistical Analysis

Statistical analyses were performed using RStudio (version 2025.05.1+153; Posit Software, Boston, MA) and R (version 4.5.1; R Foundation for Statistical Computing, Vienna, Austria). Mixed-effects modeling was conducted using the *lme4* and *ordinal* packages.

Statistically significant difference was set at *p* < .05. To examine how radiological experience modulates the benefit derived from LLM assistance, we fitted a cumulative link mixed model (CLMM), with change in top-3 accuracy (coded as -1, 0, or 1) as the ordinal outcome variable, and radiological experience in years as predictor. To investigate whether radiologist experience influenced the quality of prompts submitted to LLMs, we constructed two additional CLMMs. One model examined prompt correctness as a function of radiological experience in years, while the other assessed prompt completeness (both on a 4-point Likert scale). For all three models, random intercepts were included for both rater and case to account for individual differences among raters and variability across clinical cases.

## Code Availability

Our Python code for generating differential diagnoses via GPT-4.1, DeepSeek-R1, and Gemini 2.5 Pro is publicly available in our GitHub repository at https://github.com/shk03/expertise_paradox.

## Results

### Reader Characteristics

The neurology and neurosurgery residents did not have any formal radiological training, but a mean experience of 2.4 ± 0.5 years in their respective specialty. Radiology residents had an average of 1.6 ± 0.4 years of overall radiology experience, all of which was dedicated neuroradiology training. Neuroradiologists had 11.8 ± 4.9 years of overall radiology experience, including 6.5 ± 3.7 years in neuroradiology (Table 1).

**Table 1:** Reader characteristics. NL/NS: Neurology/Neurosurgery. RAD: Radiology.

| ID | Group | Specialty | Clinical experience overall (in years) | Radiology experience (in years) |
| --- | --- | --- | --- | --- |
| R01 | NL/NS resident | Neurology | 2.5 | 0 |
| R02 | NL/NS resident | Neurology | 2 | 0 |
| R03 | NL/NS resident | Neurosurgery | 2 | 0 |
| R04 | NL/NS resident | Neurosurgery | 3 | 0 |
| R05 | RAD resident | Radiology | 2 | 2 |
| R06 | RAD resident | Radiology | 1 | 1 |
| R07 | RAD resident | Radiology | 1.5 | 1.5 |
| R08 | RAD resident | Radiology | 3 | 1.75 |
| R09 | Neuroradiologist | Radiology | 10 | 10 |
| R10 | Neuroradiologist | Radiology | 10 | 10 |
| R11 | Neuroradiologist | Radiology | 8 | 8 |
| R12 | Neuroradiologist | Radiology | 19 | 19 |

### Diagnostic Performance

9 out of 480 case readings were excluded due to the familiarity of the readers with the case, as disclosed by the readers. Two sample cases including the case details, reader findings, and differential diagnoses are shown in Figures 2, 3.

**Figure 2:**
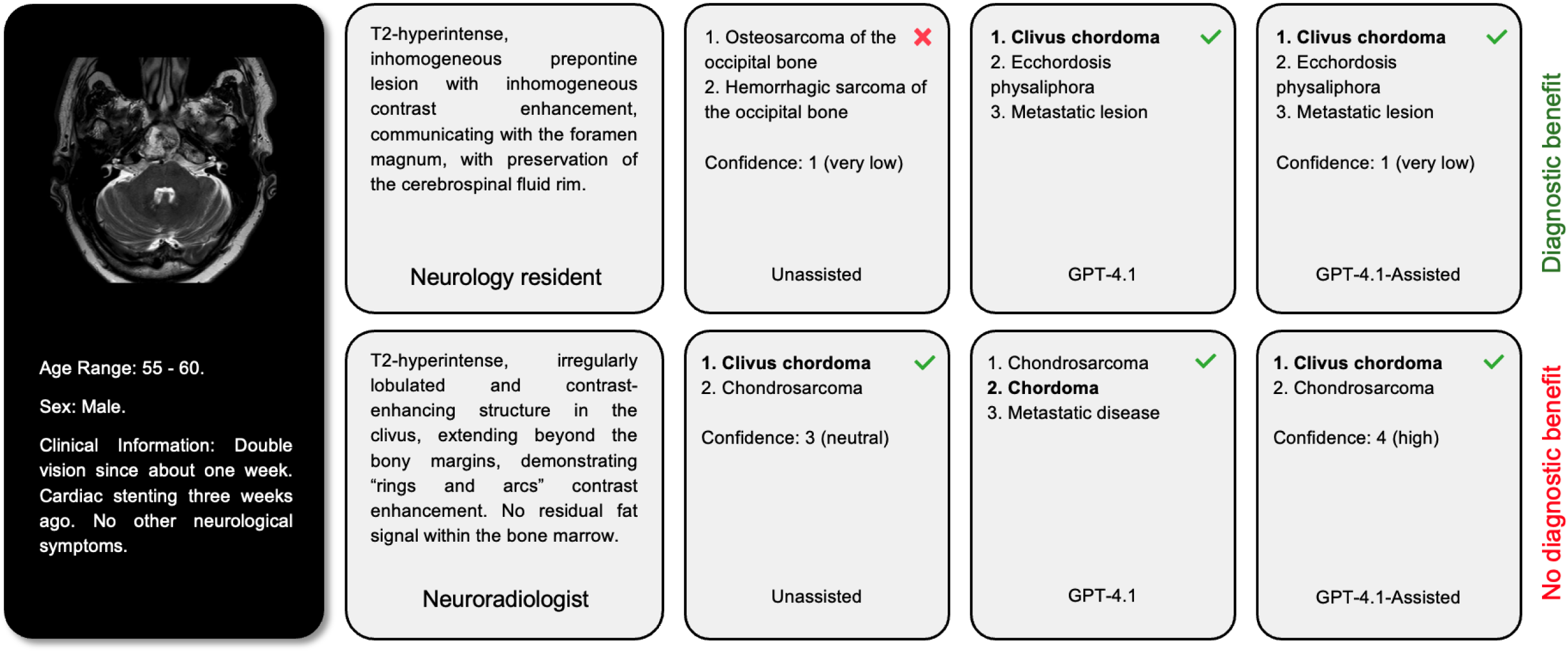
Sample case. The correct diagnosis was clivus chordoma. While GPT-4.1 suggested the correct diagnosis based on imaging findings from both the neurology resident and the neuroradiologist, only the neurology resident benefited from it, though his diagnostic confidence remained very low. Imaging descriptions were translated from German to English for illustration purposes.

**Figure 3:**
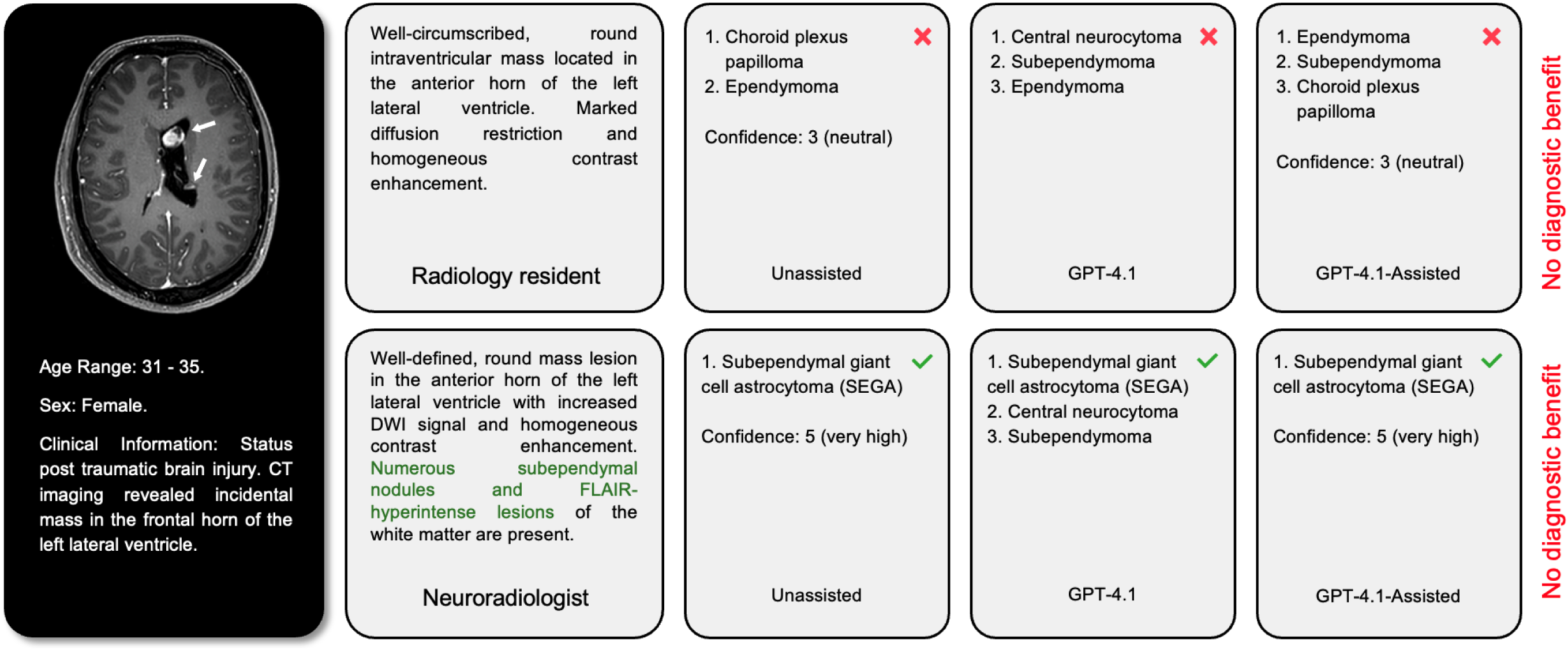
Sample case. The correct diagnosis was subependymal giant cell astrocytoma (SEGA). Although GPT-4.1 suggested the correct diagnosis based on imaging findings provided by the neuroradiologist, neither of the two readers benefited from it. Imaging descriptions were translated from German to English for illustration purposes.

For all three models, LLM-generated differential diagnoses achieved the highest top-3 accuracy when provided with image descriptions from neuroradiologists (top-3: 78.8 [126/160] - 83.8% [134/160]), followed by radiology residents (top-3: 71.8 [112/156] - 77.6% [121/156]), and NL/NS residents (top-3: 62.6 [97/155] - 64.5% [100/155]) (Figure 4). Throughout reader groups, Gemini 2.5 Pro demonstrated superior performance (NL/NS residents: 64.5% [100/155], radiology residents: 77.6% [121/156], neuroradiologists: 83.8% [134/160]).

**Figure 4:**
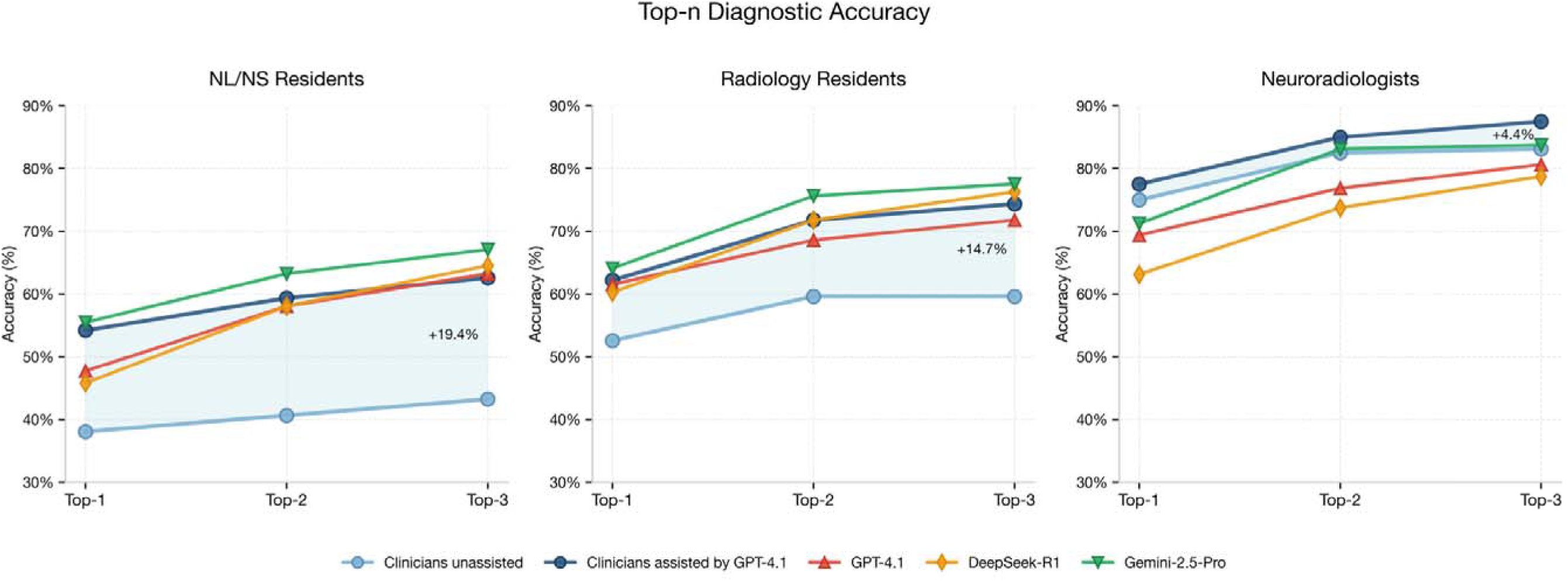
Diagnostic accuracy. The light blue area highlights the gain in diagnostic accuracy of readers through GPT-4.1 assistance. N: Neurology. NS: Neurosurgery.

Gains in top-3 accuracy from GPT-4.1 assistance diminished with increasing experience, with +19.2% for neurology/neurosurgery residents (from 43.2% [70/155] to 62.6% [97/155]),+14.7% for radiology residents (from 59.6% [93/156] to 74.4% [116/156]), and +4.4% for neuroradiologists (from 83.1% [133/160] to 87.5% [140/160]).

The CLMM revealed a significant negative association between reader experience and diagnostic benefit from LLM assistance (β = -0.098, SE = 0.035, z = -2.788, p = 0.005). This indicates that each additional year of radiological experience was associated with decreased benefit from LLM assistance. The model included random effects for both case (variance = 0.90, SD = 0.95) and rater (variance = 0.21, SD = 0.46), demonstrating substantial variation across cases and moderate variation across individual raters.

Figure 5 depicts the transitions in reader responses - from correct to incorrect and vice versa - when assisted by GPT-4.1. Across reader groups, LLM assistance resulted in a small number of correct responses being changed to incorrect, but these were outweighed by a greater number of incorrect responses corrected, yielding an overall diagnostic benefit. Only a small proportion of cases involved readers abandoning a correct diagnosis in favor of an incorrect LLM suggestion (NL/NS residents: 2.6% [4/155], radiology residents: 1.9% [3/156],neuroradiologists: 1.9% [3/160]). More commonly, readers failed to adopt correct LLM suggestions (NL/NS residents: 4.5% [7/155], radiology residents: 3.2% [5/156], neuroradiologists: 3.8% [6/160]).

**Figure 5:**
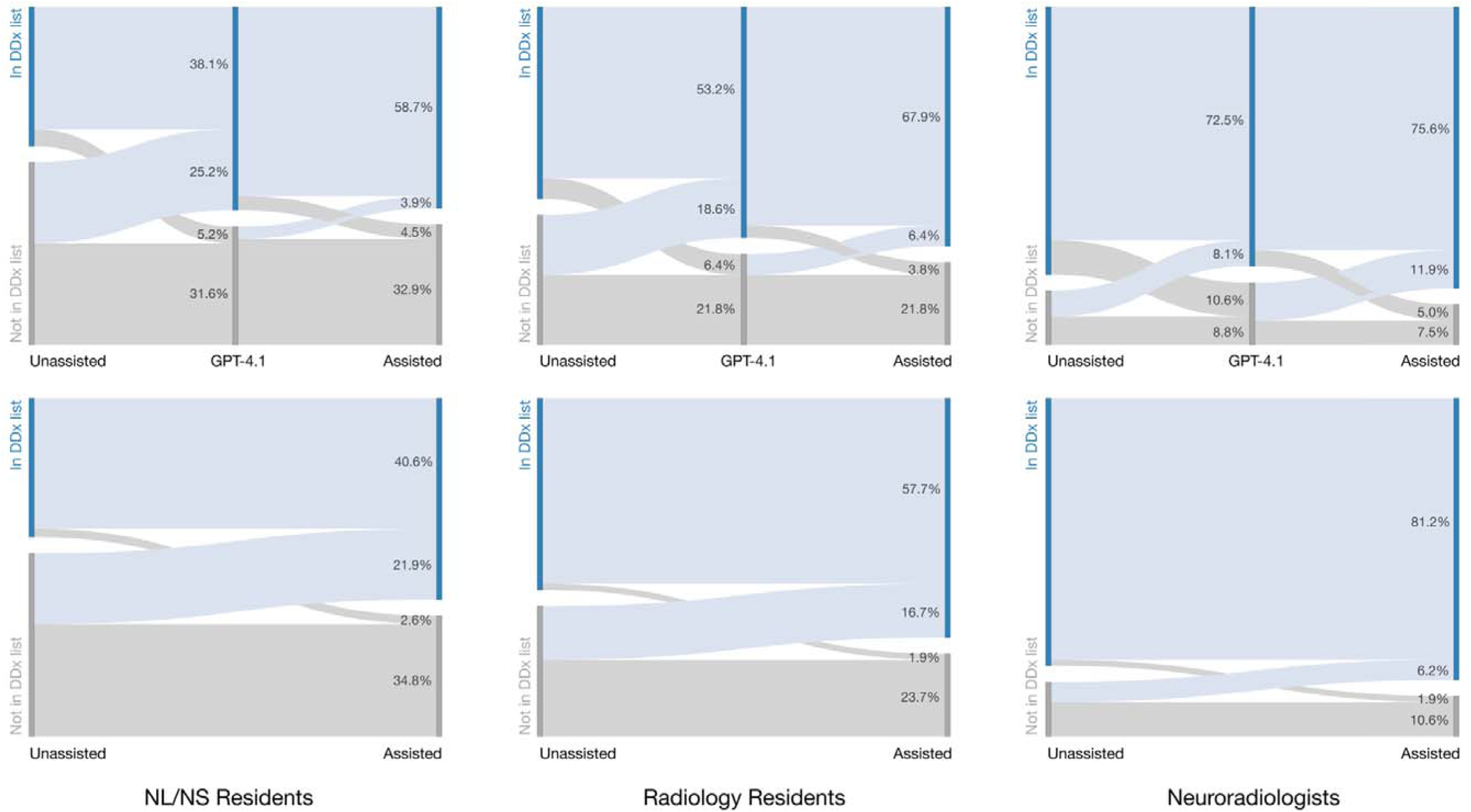
Sankey diagram illustrating the transition from correct to incorrect responses and vice versa. The first row represents the progression from unassisted readers, through GPT-4.1, to assisted readers. The second row illustrates the direct transition from unassisted to assisted readers. Throughout all reader groups, GPT-4.1 assistance led to a small number of responses changing from correct to incorrect. However, this number was smaller than the number of responses changing from incorrect to correct, resulting in an overall diagnostic benefit.

### Confidence Levels

Mean diagnostic confidence increased with GPT-4.1 assistance across all rater groups, although the magnitude of improvement diminished with increasing reader experience. Among NL/NS residents, confidence increased from 2.60 ± 1.29 to 3.41 ± 1.25. Radiology residents showed a more modest increase from 3.63 ± 1.25 to 3.88 ± 1.15, whereas neuroradiologists exhibited consistently high confidence levels, increasing only minimally from 4.41 ± 0.91 to 4.50 ± 0.96.

### Analysis of Image Descriptions

Across rater groups, imaging descriptions by neuroradiologists achieved the highest ratings for both correctness (median = 4.0, IQR = 0.0; mean = 3.70) and completeness (median = 4.0, IQR = 1.0; mean = 3.38). Radiology residents followed with high scores for correctness (median = 4.0, IQR = 1.0; mean = 3.49) but more moderate scores for completeness (median = 3.0, IQR = 2.0; mean = 2.99). NL/NS residents received the lowest ratings for both correctness (median = 4.0, IQR = 2.0; mean = 3.14) and completeness (median = 2.0, IQR = 1.0; mean = 2.27) (Figure 6).

**Figure 6:**
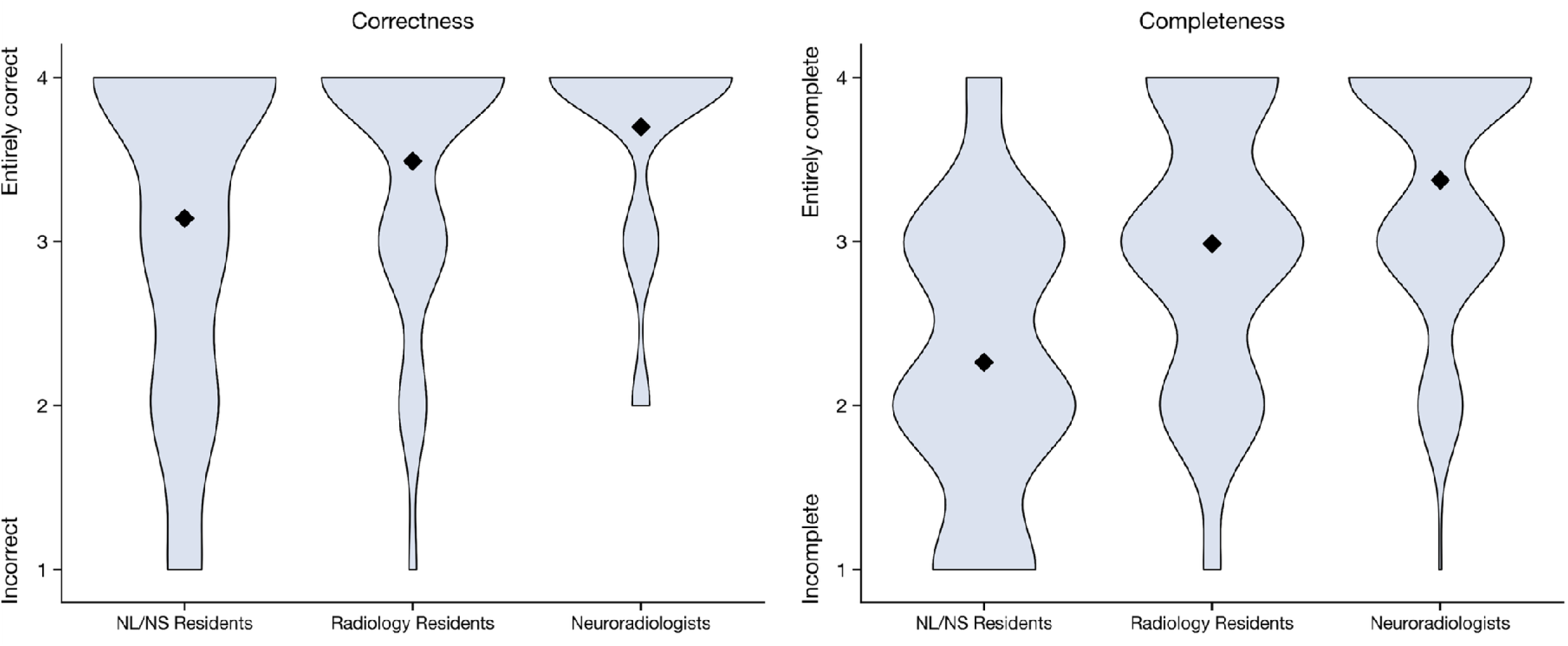
Expert ratings of reader-generated imaging findings (4-point Likert scale). Diamond symbols indicate mean scores. CLMMs revealed significant positive associations between radiological experience and both prompt correctness (β = 0.113, SE = 0.031, z = 3.714, p < 0.001) and completeness (β = 0.178, SE = 0.056, z = 3.159, p = 0.002).

CLMMs revealed significant positive associations between radiological experience and both prompt correctness (β = 0.113, SE = 0.031, z = 3.714, p < 0.001) and completeness (β = 0.178, SE = 0.056, z = 3.159, p = 0.002). Both models exhibited substantial random effects variance for case (correctness: variance = 0.89, SD = 0.94; completeness: variance = 0.93, SD = 0.96) and mixed variance for rater (correctness: variance = 0.18, SD = 0.43; completeness: variance = 1.14, SD = 1.07).

## Discussion

This study evaluated how radiologists with varying levels of experience benefit from LLM assistance in brain MRI differential diagnosis. In summary, we found that absolute performance of LLMs based on reader-generated imaging findings increased with increasing reader experience, while relative diagnostic gains of readers through LLM assistance diminished. Additionally, we validated that the differences in standalone LLM performance were associated with the correctness and completeness of imaging findings, which improved with increasing reader experience, as evaluated by an independent expert.

Our findings underline the gap between standalone LLM performance and actual clinical relevance. Even though the LLM achieved its highest performance when provided with input from expert neuroradiologists, this reader group derived only minimal benefit from LLM assistance that might not justify the effort of using such an adjunct tool. In contrast, while the present findings indicate the greatest diagnostic improvement for clinicians from other specialties without formal radiological training, this should not be interpreted as evidence that referring clinicians could use these tools to replace radiologist assessments. First, even with sizeable accuracy gains from LLM assistance, their performance remained markedly below that of neuroradiologists. Second, despite LLM support, NL/NS residents reported significantly lower confidence compared with neuroradiologists, making these assessments unreliable for clinical decision-making. Third, several critical steps in the preceding radiology workflow requiring radiologist expertise - such as protocol selection and the detection of remarkable findings - were presumed in this study, rendering an independent LLM-assisted diagnosis by referring clinicians infeasible. Considering both the practical constraints and the observed diagnostic benefits, radiology residents appear to be the group most likely to derive meaningful advantages from LLM assistance in real-world clinical practice.

Crucially, our analysis revealed substantial variation in the diagnostic benefit of LLM assistance across cases. This finding suggests that readers could harness the value of LLM-based decision support more efficiently by utilizing it in selected challenging cases.

From a broader perspective, our findings illustrate key determinants of successful human-AI collaboration in the context of generative AI. First, the quality of the content provided by the user strongly determines the quality of the output, as reflected in the standalone LLM performance by reader group. Unlike traditional machine learning systems executing narrowly defined tasks on fixed input data, generative AI systems can perform a much broader range of tasks. Yet, performance is highly contingent on the clarity, precision, and contextual richness of the model prompt. One study reported improvements in diagnostic performance by up to 30% when incorporating lab results in the LLM prompt [18]. Alarmingly, however, LLMs are also vulnerable to cognitive biases, such as suggestibility bias (prioritizing user agreement over independent reasoning) and framing bias (influence of presentation or wording on decision-making) [19]. For example, salient but distracting patient history information was shown to impair LLM diagnostic performance [20]. Second, the capacity of the human user to differentiate between correct and incorrect LLM suggestions is essential. This ability is shaped by the relative degree of trust placed in one’s own judgment versus trust in the model’s outputs, as suggested by prior research on automation bias (overreliance on automated decision-making systems) and algorithmic aversion (excessive skepticism toward algorithmic outputs) [21–24]. Interestingly, a non–domain-specific study on user perceptions of LLM accuracy found that longer explanations increased users’ confidence in the LLM’s responses, even when the actual response accuracy remained unchanged [25]. In clinical practice, where physicians face enormous workload pressures, cognitive and temporal constraints further limit the users’ capacity to verify the large volumes of content that LLMs can generate [26]. There are ongoing research efforts to explore methods for quantifying the certainty of LLM outputs, and early findings suggest that token-level probability estimates may provide a more reliable indicator of model uncertainty than self-reported confidence scores, which tend to exhibit overconfidence [27]. However, the mechanisms by which trust in LLMs is formed, maintained, or undermined within clinical decision support contexts remain largely underexplored, representing a promising avenue for future research.

### Limitations

This study has several limitations. First, this is a single-center study employing a small set of cases in a sub-domain of radiology, limiting the generalizability of findings. Second, we deliberately standardized the interaction of readers with LLMs to avoid the introduction of undesired confounders, but this reduced the realism of the setting. Variations in prompting strategies, choice of models or tools, and the amount of effort devoted to verifying LLM suggestions will likely amplify differences in reader-level benefits. Third, the imaging findings were presented to the models in German, the native language of the readers. Prior studies have shown that LLMs tend to achieve higher performance when prompted in high-resource languages [28], and providing the inputs in English might have resulted in slightly better model performance. Finally, the models in this study were provided with textual information only. This decision was based on prior evidence indicating only negligible accuracy improvements when including MRI key images as additional input [17]. Recently, vision-language models (VLM) able to process 3D medical imaging data have been described, but their availability remains limited [29, 30]. The diagnostic benefits of human readers from such models remain to be investigated.

## Conclusion

With increasing reader experience, the absolute diagnostic performance of LLMs in brain MRI cases based on reader-generated imaging findings improved, while the relative diagnostic benefit of LLM assistance declined. Our findings call attention to the gap between standalone LLM performance and actual clinical relevance, emphasizing the need to account for human-AI interaction in this context.

## Data Availability

All data produced in the present study are available upon reasonable request to the authors

---

**Supplement 1:**
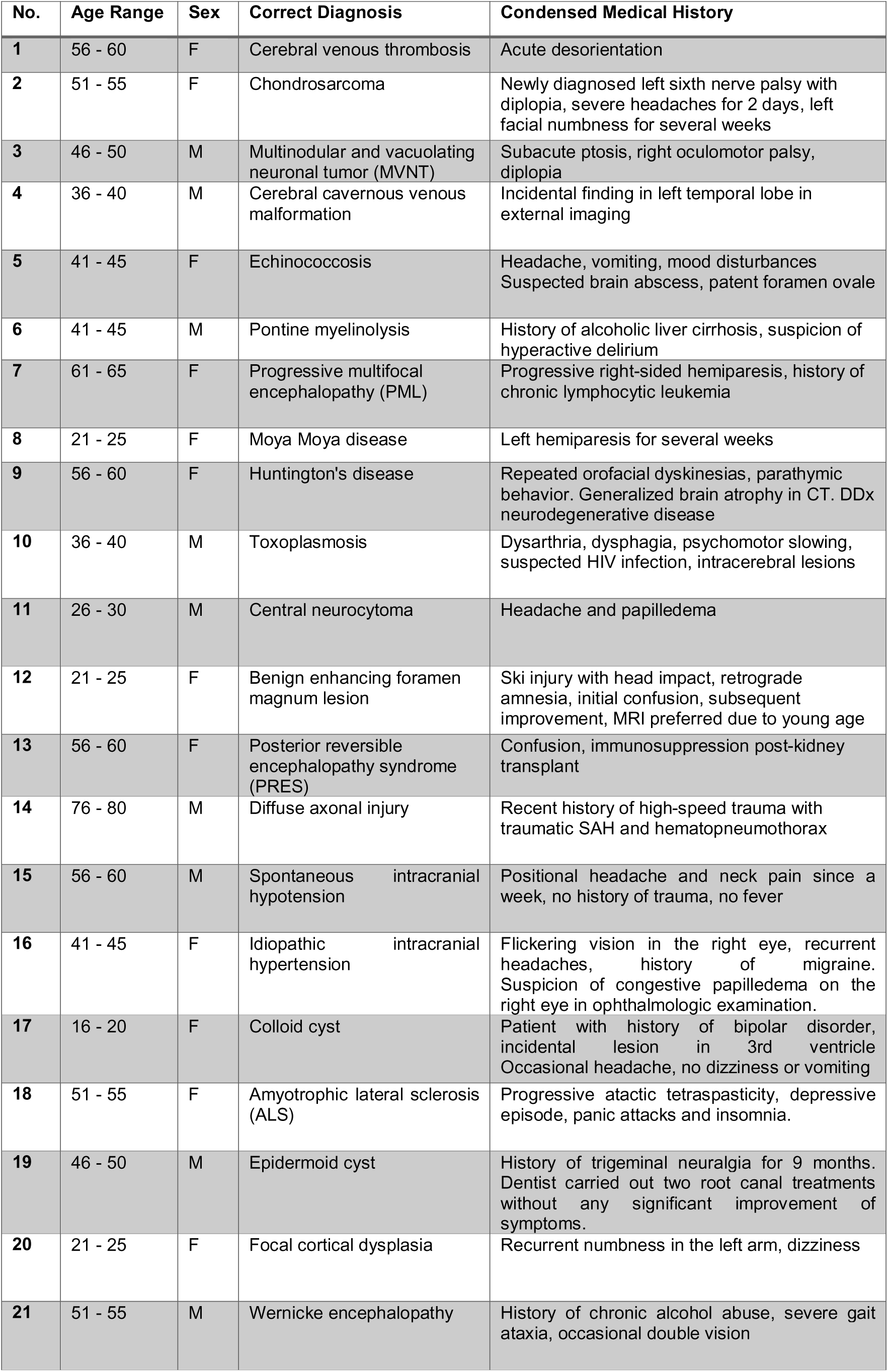
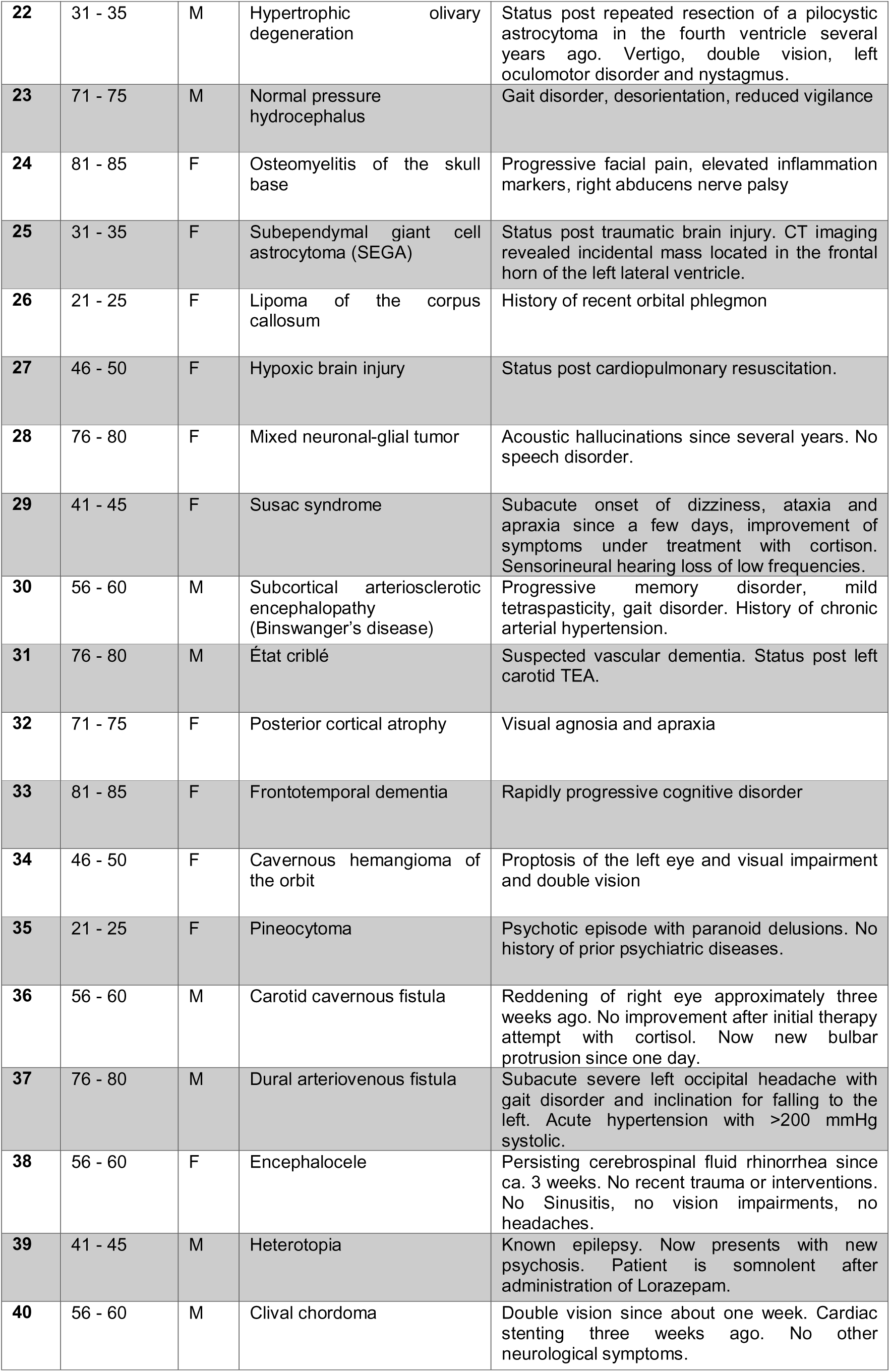
Case overview.

**Supplement 2:** Adjudication of edge cases.

| Correct Diagnosis | Reader / LLM Response | Consensus Decision |
| --- | --- | --- |
| Toxoplasmosis | Parasitic infection | Incorrect |
| Toxoplasmosis | HIV-associated opportunistic infection | Incorrect |
| CSF leak | Bilateral hygroma | Incorrect |
| Subcortical arteriosclerotic encephalopathy (Binswanger disease) | Hypertensive leukoencephalopathy | Correct |
| Subcortical arteriosclerotic encephalopathy (Binswanger disease) | Cerebral small vessel disease | Correct |
| Osteomyelitis of the skull base | Mastoiditis | Incorrect |
| Mixed neuronal-glial tumor | Dysembryoplastic Neuroepithelial Tumor (DNET) | Correct |
| Mixed neuronal-glial tumor | Ganglioglioma | Correct |
| Multinodular and vacuolating neuronal tumor (MVNT) | Mixed glioneuronal tumor | Incorrect |
| Pineocytoma | Pinealoma | Correct |
| Cavernous hemangioma of the orbit | Retroorbital low-flow malformation | Correct |

**Supplement 3:** Legend for expert ratings of reader-generated imaging findings (4-point Likert scale).

*Supplement 2: Adjudication of edge cases.*
| **Correctness** |  |  |
| --- | --- | --- |
| 1 | Incorrect | Several major errors are present that undermine the diagnostic interpretation. |
| 2 | Partially Correct | At least one major error is present that undermines the diagnostic interpretation. |
| 3 | Mostly Correct | Findings are generally accurate, with only minor inaccuracies that do not undermine the diagnostic interpretation. |
| 4 | Entirely Correct | All documented findings and interpretations are accurate. No errors. |
| **Completeness** |  |  |
| 1 | Incomplete | Several key findings are omitted, undermining the diagnostic interpretation. |
| 2 | Partially Complete | At least one key finding is omitted, undermining the diagnostic interpretation. |
| 3 | Mostly Complete | Most relevant findings are documented, with only minor omissions. |
| 4 | Entirely Complete | All relevant findings are thoroughly documented. No omissions relevant to the diagnosis. |

## Notes

### Competing Interest Statement

The authors have declared no competing interest.

### Funding Statement

This study did not receive any funding

### Author Declarations

Ethics Committee of the Technical University of Munich gave ethical approval for this work

